# Dynamic Shifts in the Oral Microbiota Following Cancer Surgery: A 172-Sample Longitudinal Study of Surgical Site Infection Risk

**DOI:** 10.64898/2026.05.18.26353519

**Authors:** Marianna Sampaio Serpa, Alexandre Defelicibus, Thais F. Bartelli, Israel Tojal da Silva, Diana Noronha Nunes, Luiz Paulo Kowalski, Emmanuel Dias-Neto

## Abstract

**Background:** Surgical site infection (SSI) is the leading cause of perioperative morbidity following oral cancer surgery, yet the role of the oral microbiota in SSI pathogenesis remains poorly defined. This study prospectively investigated microbiota dynamics in relation to SSI occurrence in patients undergoing resection for oral squamous cell carcinoma (OSCC).

**Methods:** A total of 172 oral swab samples were collected from 45 OSCC patients across four longitudinal time points: baseline (∼29 days pre-surgery), immediately pre-surgery (hospital admission), early post-surgery (within 5 days), and late post-surgery (6–15 days). Bacterial composition was profiled by 16S-rDNA V3-V4 sequencing (172 successfully sequenced samples), and bacterial/human DNA ratios were quantified by qRT-PCR (170 samples evaluated). SSI was assessed within 30 days post-surgery using adapted CDC criteria.

**Results:** Fourteen of 45 patients (31.1%) developed SSI. Younger age was significantly associated with SSI occurrence (median age 53.2 years in SSI group vs. 67.4 years in non-SSI group; p=0.011), with each one-year decrease in age conferring a 7% increased risk. Notably, younger patients presented with larger and more advanced tumors (T3/T4: median age 57.2 vs. 72.9 years for T1/T2; p=0.033), leading to more extensive surgical procedures. Across all 172 samples, surgery induced a marked post-operative reduction in bacterial load and diversity. However, at the late post-surgery time point (collection IV), patients with SSI exhibited significantly higher alpha-diversity compared to non-infected patients (p<0.05 for Observed, Shannon, and Simpson indices). Beta-diversity also differed significantly between groups at this time point (weighted UniFrac, p=0.043). *Prevotella* and *Porphyromonas* dominated SSI patients at infection, together accounting for ∼40% of reads versus 9.5% in non-infected patients. Among the 172 samples analyzed longitudinally, *Aggregatibacter* abundance at the early post-surgery time point (collection III) emerged as a significant predictor of subsequent SSI (OR per 1% increase: 1.10; p=0.012), with frequencies >0.044% conferring a 5.7-fold higher risk.

**Conclusions:** Our longitudinal analysis demonstrate that while OSCC surgery profoundly disrupts the oral microbiota, non-SSI patients restore their preoperative profile within 12 days. In contrast, SSI is characterized by persistent dysbiosis dominated by *Prevotella* and *Porphyromonas*. Younger patients with advanced tumors are at particular risk. Early post-surgical *Aggregatibacter* abundance may serve as a novel risk indicator for SSI, potentially enabling timely preventive interventions in high-risk patients.

## Introduction

Oral cancer (OC), primarily squamous cell carcinoma arising from the oral cavity (lips, tongue, floor of mouth, gingiva, buccal mucosa, palate), represents a significant global health burden, with approximately 390,000 new cases and 188,000 deaths annually worldwide (Globocan, 2022). It disproportionately affects regions with high tobacco and alcohol use, and early detection is critical as 5⍰year survival drops from >80% in early stages to <40% in advanced disease (Dhingra, 2025). Treatment is multimodal but surgery remains the cornerstone, offering the best chance for cure in nonmetastatic cases through transoral resection or more extensive composite resections with neck dissection, followed by adjuvant radiotherapy ± chemotherapy based on risk factors (Colevas et al., 2025).

Despite these advances, oncological treatments for OC—particularly surgery—carry substantial risks of complications. Surgical management of OC can lead to several postoperative issues, including surgical site infection (SSI), respiratory infections, flap necrosis, sepsis, and even death (Jain et al., 2025; Mak et al., 2025). Among these, SSI is the leading cause of perioperative morbidity after head and neck cancer surgery. Recent meta-analyses have systematically quantified the burden of SSI in oral cancer surgery. Qin et al. (2026) pooled data from 21 studies (6,071 patients) and reported an overall SSI incidence of 29% (95% CI: 25%–33%), with higher rates observed in advanced-stage disease (34%), T3/T4 tumors (42%), bilateral neck dissection (48%), and free flap reconstruction (32%). Similarly, Chen et al. (2025) analyzed 9 studies (4,360 participants) and found a pooled SSI incidence of 22.22%, identifying free flap reconstruction (OR 4.50), mandibulectomy (OR 4.35), diabetes mellitus (OR 3.67), and perioperative blood transfusion (OR 3.04) as significant risk factors. Together, these meta-analyses confirm that SSI affects approximately one-quarter to one-third of oral cancer surgical patients, with risk substantially increased by procedural complexity and host factors.

Defined as an infection at the surgical site within 30 days postoperatively (Holihan et al., 2017), SSI in oral cancer patients results in profound consequences including significant functional deficits such as speech and swallowing impairment (Kolokythas et al., 2010; Nichols et al., 2025), prolonged antibiotic use (Qin et al., 2026), extended hospital stays with escalated costs (Fan et al., 2025; Balagopal et al., 2022), and higher mortality (Balagopal et al., 2022; Qin et al., 2026). Moreover, SSI delays the initiation of essential adjuvant therapies, such as radiotherapy and chemotherapy, which are critical for high-risk patients (Qin et al., 2026). Notably, Grandis et al. (1992) demonstrated that patients who develop SSI face a significantly higher risk of locoregional recurrence compared to those without postoperative infection.

Emerging evidence highlights the potential role of the oral and surgical site microbiota—particularly pathogenic bacteria—in driving SSI pathogenesis through dysbiosis, biofilm formation, and opportunistic overgrowth in the perioperative period (Santacroce et al., 2025; Huang et al., 2025). The oral cavity serves as a reservoir for a diverse array of microorganisms, and alterations in its microbial composition have been linked to local inflammation, immune responses, and compromised wound healing following surgical intervention (Santacroce et al., 2025). Postoperative head and neck cancer patients frequently experience oral microbiome dysbiosis due to surgical trauma and reduced oral function, which may exacerbate symptoms and impair recovery (Huang et al., 2025). In this study, we investigated the longitudinal microbiota dynamics in OC patients before and after surgery, including a cohort of cases that developed SSI, to elucidate microbial shifts associated with postoperative infectious complications.

## Patients, Materials and Methods

The study protocol was submitted to and approved by the institutional Research Ethics Committee of the A.C. Camargo Cancer Center prior to the collection of the first sample (Protocol No. 2188/2016). Between 2016 and 2019, a total of 68 patients diagnosed with oral squamous cell carcinoma (OSCC) were invited to participate in this prospective study. Of these, 45 patients underwent tumor excision surgery at the A.C. Camargo Cancer Center and were followed from diagnosis until ∼30 days after surgery, and included in the final cohort for analysis of the oral microbiota and surgical site infection (SSI) occurrence. All participants provided written informed consent before enrollment.

To gather clinical information, questionnaires and medical charts were used. Additionally, an oral clinical exam was conducted to assess the dental and periodontal health of the patients. The dental conditions were evaluated by calculating the sum of decayed, missing, and filled teeth (DMFT) (Klein et al., 1938), while the periodontal health was evaluated by measuring the probing depth (PD) - the distance from the gingival margin to the bottom of the periodontal pocket - and gingival recession (GR) - the distance from the gingival margin to the cemento-enamel junction (Lindhe & Nyman, 2005). By adding the PD and GR measurements, the clinical attachment level (CAL) was calculated to indicate the total attachment loss of the periodontium.

Presence of SSI was assessed based on an adaptation of the criteria described by the Center for Disease Control and Prevention (CDC, 2014) when any of the following events occurred: the presence of purulent material in the patient’s incision or drain, or a diagnosis of SSI by the physician (surgeon or infectious disease specialist), which was usually based on the presence of spontaneous wound dehiscence with signs or symptoms of SSI (pain, erythema, redness, and/or swelling in the operated area, fever, or an increase in white blood cell count). Infection accompanied by the development of an orocutaneous fistula was also considered an SSI. The presence of SSI was evaluated within 30 days after surgery.

### Sample collection

In total, 172 samples were collected from the 45 subjects to allow microbiome assessment in 4 different moments: I - prior to surgery (baseline), when patients were undergoing preoperative exams; II - just before the surgery (when hospitalized); III - within 5 days after surgery; IV - between 6-15 days after surgery (**Figure 1**).

**Figure 1.**
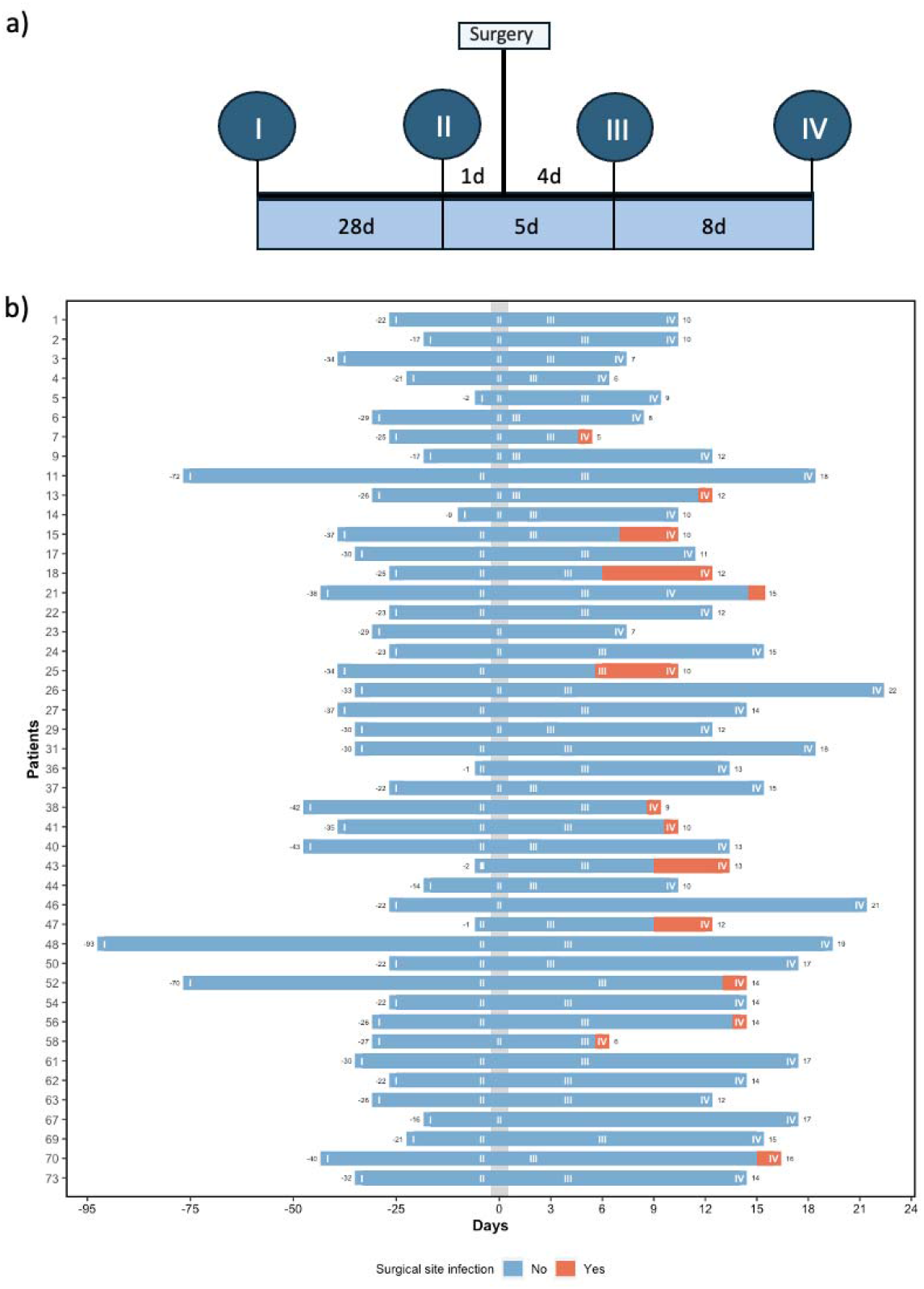
Collection points and events of surgical site infections (SSIs) in the patients enrolled in the study. Panel A shows the average time (in days) between the 4 collection points and surgery for tumor excision. Panel B shows the exact number of days for each patient. Occurrence of SSI is marked with orange boxes for the 14 SSI-positive individuals.

The samples were collected using FLOQSwab 519C oral swabs (COPAN *Diagnostics*, USA). Before surgery a swab was taken from the area adjacent to the tumor, and after surgical excision a sample was collected from the area near the surgical wounds. Following collection, the swabs were placed back into their respective tubes and 2ml of a stabilizing solution (NaCl 0.1M; TrisHCl 0.01M, pH 7.5; EDTA 0.01M; 0.5% SDS and proteinase K 0.1mg/ml) was added to cover the tip of each swab (Min et al., 2006). All tubes were vortexed for 10 seconds and stored in a refrigerator until DNA extraction using the DNeasy PowerSoil Kit 12888 (MoBio. Qiagen. USA). DNA samples were quantified using the Qubit dsDNA High Sensitivity kit (Invitrogen, Carlsbad, USA).

### Real time PCR: Quantitative analysis of bacteria and human DNA

Human:bacteria DNA-ratios were determined in our samples by quantitative real-time PCR (qRT-PCR) as described (de Albuquerque et al., 2002). For this, two pairs of primers were used: one targeting a small conserved internal region of the V1 16S-rDNA gene (forward: 5’-AGAGTTTGATCMTGGCTCAG-3’; reverse: 5’-TTACTCACCCGTICGCCRCT-3’) and another targeting a fragment of the human beta-actin gene (forward: 5’-CCATCTACGAGGGGTATGC-3’; reverse: 5’-GGTGAGGATCTTCATGAGGTA-3’). These primer-pairs generate amplicons of 113bp (16S-V1) and 90bp (beta-actin). Amplification reactions were performed separately, using the same amount of DNA (5ng), 2µM (V1 16S-rDNA) or 5µM (human beta-actin) of each primer-pair, and the Fast SYBRTM Green Master Mix enzyme (ThermoFisher Scientific, Massachusetts, USA). Amplifications were performed using the 7500 Fast platform (Thermo Fisher Scientific, Massachusetts, USA), starting with a denaturation step at 95°C for 20s, followed by 40 cycles of 95°C for 3s and 60°C for 30s.

### Bacterial 16S-rDNA gene sequencing and bioinformatic analysis

#### Target gene amplification (16S-rDNA) and sequencing

Bacterial content was assessed in this study through the amplification and sequencing of the V3-V4 regions of the 16S ribosomal DNA (rDNA) gene. Amplification primers were designed for the Illumina platform sequencing and generated amplicons with an average size of 510 bp. Primer sequences were U341F (5’-CACTCTTTCCCTACACGACGCTCTTCCGATCTCCTACGGGRSGCAGCAG-3’), and 806R (5’-GTGACTGGAGTTCAGACGTGTGCTCTTCCGATCTGGACTACHVGGGTWTCTAAT-3’), both including adapters suitable for Illumina platform sequencing. The amplification was performed with 16ng of DNA, 2.5µM of each primer, and KAPA HiFi HotStart ReadyMix (2X) enzyme (Kapa Biosystems. Sigma Aldrich. USA). The cycling conditions were 95°C for 2min, followed by 95°C for 20s, and 54°C for 15s (28 cycles), followed by a final extension step at 72°C for 5min. The amplicons were purified with Ampure XP Beads and quantified in Qubit dsDNA High Sensitivity. The libraries were constructed and sequenced by Neoprospecta Microbiome Technologies using the MiSeq Reagent v2 kit (500-cycles) from Illumina, paired-end (300nt read 1 and 210nt read 2) on the MiSeq platform, aiming to generate ≥ 50.000 reads per sample (Christoff et al., 2017).

#### Processing and filtering sequenced samples

The data processing and filtering steps included: 1-removal of adapters and primers; 2-removal of reads that mapped with the human genome; 3-import of the reads to the program Quantitative insights into microbial ecology 2 (Qiime2); 4-junction of paired reads and quality filter (phred score ≥10); 5-removal of noise from reads through deblur. The sequences were clustered as Operational Taxonomic Units (OTUs) when they presented 97% identity between them, followed by taxonomic classification of the OTUs using the SILVA database (Quast et al., 2012). Rarefaction curves were used to ensure that all the samples were saturated and could be analyzed, OTUs with less than three reads were discarded to remove potential remaining sequencing errors. Analyses performed here were limited to the genus level.

## Statistical analysis

Research Electronic Data Capture (REDCap) system recorded all clinical data. To optimize analysis, several variables were categorized, including skin color (white x non-white), smoking habit (non-smokers or former smokers x smokers), alcohol drinking habit (non-alcohol-drinkers or former drinkers x alcohol-drinkers), tumor size (T1/T2 x T3/T4), and clinical stage (I/II x III/IV). The information related to clinical variables and RT-PCR results were transferred to the Statistical Package for the Social Sciences (SPSS) version 17.0 (SPSS Inc., Chicago, IL, USA) for further analysis. Fisher and Chi-square tests were employed to evaluate categorical data, while the Mann-Whitney U test was used for quantitative data. The Friedman test was utilized to analyze the relationship between the mean cycle threshold (CT) of RT-PCR and collection time-point of SSI. In addition, to investigate the association between SSI, CT, and the collection point, the Mann-Whitney U test was utilized.

The analysis of metagenomic data was conducted using the phyloseq package in RStudio program (version 3.5.1; RStudio: Integrated Development for R. RStudio Inc., Boston, MA, USA). The Mann-Whitney U test was used to determine statistically significant differences between SSI and alpha-diversity and multivariate Analysis of Similarity test (ANOSIM) was employed to evaluate beta diversity. Differences in microbial composition between groups were determined by the Linear discriminant analysis Effect Size (LEfSe) software, based on bacterial frequency and LDA score generated by the program (Segata et al., 2011). A simple logistic regression model was used to assess the likelihood of developing SSI based on clinical data, RT-PCR, and sequencing results (bacterial diversity and frequency). All statistical tests were conducted at a significance level of 5% (p<0.05).

## Results

### Clinical data

A total of 51 patients were initially enrolled in this study. However, six were excluded because their surgeries were canceled due to: i) detection of metastatic lung disease (n = 3); ii) voluntary withdrawal from surgery (n = 2); and iii) observation of carotid artery involvement by the tumor (n = 1). This resulted in a final cohort of 45 subjects. Most of the patients were male (n=32; 71.1%) with a mean age of 63 years. Among smokers/ex-smokers (60%), the average consumption was 25 cigarettes per day for 38 years. Regarding alcohol use, 73.4% were alcoholics/ex-alcoholics, with an average consumption time of 38 years. The most frequent comorbidities were hypertension (n=25; 55.6%) and chronic obstructive pulmonary disease (COPD) (n=13; 28.9%). Concerning the tumor, based on clinical and imaging evaluation, the tongue was identified as the most common primary site (n=20; 44.4%), followed by the floor of the mouth (n=9; 20%), buccal/retromolar region (n=7; 15.6%), inferior lip (n=6; 13.3%), hard palate (n=2; 4.5%) and gingiva (n=1; 2.2%). Most tumors were categorized as clinical stage III/IV (n=33; 76.7%). Concerning oral examination, 7 patients (15.6%) were edentulous, and for the remaining 38 patients (84.4%), the mean number of teeth was 18. The mean DMFT score was 20.5 and mean pocket depth was 2.8mm. Most patients who underwent lymph node dissection were submitted to a bilateral drainage extension (n=20; 55.5%), while 44.4% underwent ipsilateral dissection (n=16). The average surgery time was 9 hours, and the intensive care unit (ICU) hospitalization had an average of 3.6 days. Fourteen patients (14/45: 31.1%) developed SSI.

In this cohort, SSI was more frequent in younger patients, in those with more advanced clinical stages III/IV, and in patients that underwent cervical dissection and surgical reconstruction, as well as in those with longer operating time (p<0.05). Logistic regression analysis showed that patients with larger tumors (T3/T4) and advanced clinical stage (III/IV) have 6.5 and 9.3 times more chances of developing SSI as compared to patients with smaller tumors and stage I/II (**Table 1**). Regarding age, each one-year decrease in age was associated with a 7% increase in the risk of developing SSI.

**Table 1.** Clinical variables and SSI.

| Variables | n | Surgical site infection |  | p-value<sup>1</sup> | OR | CI 95% | p-value<sup>2</sup> |
| --- | --- | --- | --- | --- | --- | --- | --- |
|  |  | No | Yes |  |  |  |  |
| Qualitative variables |  |  |  |  |  |  |  |
| Sex |  |  |  |  |  |  |  |
| Male | 32 | 22 (71.0%) | 10 (71.4%) | 0.999<sup>a</sup> | 0.98 | 0.24-3.94 | 0.975 |
| Female | 13 | 9 (29.0%) | 4 (28.6%) |  |  |  |  |
| Skin color |  |  |  |  |  |  |  |
| White | 26 | 18 (58.1%) | 8 (57.1%) | 0.999<sup>b</sup> | 1.04 | 0.28-3.72 | 0.954 |
| Non-white | 18 | 13 (41.9%) | 6 (42.9%) |  |  |  |  |
| Tobacco consumption |  |  |  |  |  |  |  |
| No/Former smoker | 3<sup>3</sup> | 12 (70.6%) | 21 (80.8%) | 0.481<sup>b</sup> | 0.57 | 0.14-2.38 | 0.442 |
| Yes | 0<sup>1</sup> | 5 (29.4%) | 5 (19.2%) |  |  |  |  |
| Alcohol consumption |  |  |  |  |  |  |  |
| No/Former drinker | 8<sup>1</sup> | 9 (52.9%) | 9 (34.6%) | 0.382<sup>a</sup> | 2.12 | 0.61-7.41 | 0.237 |
| Yes | 5<sup>2</sup> | 8 (47.1%) | 17 (65.4%) |  |  |  |  |
| Comorbidity |  |  |  |  |  |  |  |
| No | 9 | 5 (16.1%) | 4 (28.6%) | 0.428<sup>a</sup> | 0.481 | 0.11-2.16 | 0.340 |
| Yes | 36 | 26 (83.9%) | 10 (71.4%) |  |  |  |  |
| Arterial hypertension |  |  |  |  |  |  |  |
| No | 20 | 13 (41.9%) | 7 (50.0%) | 0.857<sup>b</sup> | 0.72 | 0.20-2.57 | 0.615 |
| Yes | 25 | 18 (58.1%) | 7 (50.0%) |  |  |  |  |
| COPD |  |  |  |  |  |  |  |
| No | 20 | 22 (71.0%) | 10 (71.4.0%) | 0.999<sup>a</sup> | 0.98 | 0.24-3.94 | 0.975 |
| Yes | 25 | 9 (29.0%) | 4 (28.6%) |  |  |  |  |
| Tumor size<sup>c</sup> |  |  |  |  |  |  |  |
| T1/T2 | 18 | 16 (59.3%) | 2 (18.2%) | 0.052<sup>b</sup> | 6.54 | 1.18-36.32 | 0.032<sup>*</sup> |
| T3/T4 | 20 | 11 (40.7%) | 9 (81.8%) |  |  |  |  |
| Lymph node metastasis<sup>†</sup> |  |  |  |  |  |  |  |
| No | 1<sup>2</sup> | 17 (63%) | 4 (36.4%) | 0.167<sup>a</sup> | 2.98 | 0.69-12.76 | 0.142 |
| Yes | 7<sup>1</sup> | 10 (37%) | 7 (63.6%) |  |  |  |  |
| Clinical stage<sup>†</sup> |  |  |  |  |  |  |  |
| I/II | 10 | 13 (48.1%) | 1 (9.1%) | 0.030<sup>a</sup> | 9.29 | 1.04-82.96 | 0.046<sup>*</sup> |
| III/IV | 33 | 14 (51.9%) | 10 (90.9%) |  |  |  |  |
| ASA |  |  |  |  |  |  |  |
| I/II | 32 | 23 (74.2%) | 9 (64.3%) | 0.502<sup>a</sup> | 1.60 | 1.41-6.21 | 0.499 |
| III/IV | 13 | 8 (25.8%) | 5 (35.7%) |  |  |  |  |
| Type of tissue removed |  |  |  |  |  |  |  |
| Soft | 24 | 17 (54.8%) | 7 (50.0%) | 0.999<sup>b</sup> | 1.21 | 0.34-4.30 | 0.763 |
| Soft and hard | 21 | 14 (45.2%) | 7 (50.0%) |  |  |  |  |
| Cervical drainage |  |  |  |  |  |  |  |
| No | 9 | 9 (29.0%) | 0 (0.0%) | 0.040<sup>a*</sup> | NA |  |  |
| Yes | 36 | 22 (71.0%) | 14 (100.0%) |  |  |  |  |
| Cervical drainage extension |  |  |  |  |  |  |  |
| Ipsilateral | 16 | 12 (54.5%) | 4 (28.6%) | 0.236<sup>b</sup> | 3.00 | 0.72-12.56 | 0.132 |
| Bilateral | 20 | 10 (45.5%) | 10 (71.4%) |  |  |  |  |
| Surgical reconstruction |  |  |  |  |  |  |  |
| No | 13 | 13 (41.9%) | 0 (0.0%) | 0.004<sup>a*</sup> | NA |  |  |
| Yes | 32 | 18 (58.1%) | 14 (100.0%) |  |  |  |  |
| Tracheostomy |  |  |  |  |  |  |  |
| No | 16 | 12 (38.7%) | 2 (14.3%) | 0.165<sup>a</sup> | 3.79 | 0.72-19.98 | 0.116 |
| Yes | 20 | 19 (61.3%) | 12 (85.7%) |  |  |  |  |
| Prophylactic antibiotic<sup>†</sup> |  |  |  |  |  |  |  |
| Clindamycin and Gentamicin | 18 | 16 (55.2%) | 12 (85.7%) | 0.104<sup>b</sup> | 0.205 | 0.04-1.09 | 0.062 |
| Other | 15 | 13 (44.8%) | 2 (14.3%) |  |  |  |  |
| Prophylactic mouth wash<sup>†</sup> |  |  |  |  |  |  |  |
| No | 29 | 22 (71.0%) | 7 (53.8%) | 0.313<sup>a</sup> | 2.01 | 0.55-7.99 | 0.279 |
| Yes | 15 | 9 (29.0%) | 6 (46.2%) |  |  |  |  |
| **Intraoperative transfusion** |  |  |  |  |  |  |  |
| No | 38 | 28 (90.3%) | 10 (71.4%) | 0.180<sup>a</sup> | 3.73 | 0.71-19.67 | 0.120 |
| Yes | 7 | 3 (9.7%) | 4 (8.6%) |  |  |  |  |
| **ICU** |  |  |  |  |  |  |  |
| No | 11 | 10 (32.3%) | 1 (7.1%) | 0.132<sup>b</sup> | 6.19 | 0.71-54.16 | 0.099 |
| Yes | 34 | 21 (67.7%) | 13 (92.9%) |  |  |  |  |
| **Quantitative variables** |  |  |  |  |  |  |  |
| **Age** |  |  |  |  |  |  |  |
| Mean (SD) |  | 66.6 (12.9) | 54.9 (12.7) | 0.011<sup>c</sup> | 0.93 | 0.88-0.99 | 0.016* |
| Median (min-max) |  | 67.4 (39.3-88.7) | 53.2 (25.9-76.3) |  |  |  |  |
| **BMI** |  |  |  |  |  |  |  |
| Mean (SD) |  | 26 (4.3) | 23.3 (4.2) | 0.050<sup>c</sup> | 0.85 | 0.71-1.00 | 0.057 |
| Median (min-max) |  | 25.8 (17.5-37.9) | 22.6 (16.9-31.1) |  |  |  |  |
| **DMFT** |  |  |  |  |  |  |  |
| Mean (SD) |  | 21.7 (5.6) | 17.93 (7.7) | 0.135<sup>c</sup> | 0.92 | 0.79-1.03 | 0.125 |
| Median (min-max) |  | 24 (11-28) | 19 (12-28) |  |  |  |  |
| **NIC** |  |  |  |  |  |  |  |
| Mean (SD) |  | 2.6 (1.3) | 3.3 (2.3) |  |  |  | 0.117 |
| Median (min-max) |  | 2.3 (1-6.4) | 1.8 (1.2-9.3) | 0.548<sup>c</sup> | 1.47 | 0.91-2.39 |  |
| **Surgical time (hours)** |  |  |  |  |  |  |  |
| Mean (SD) |  | 8.2 (5.4) | 11.3 (3.6) | 0.048<sup>c*</sup> | 1.14 | 0.10-1.31 | 0.057 |
| Median (min-max) |  | 9.2 (1.3-18.0) | 10.9 (6.33-17.0) |  |  |  |  |

1: Independence test; 2: Simple logistic regression model; a: Chi-square de Pearson test; b: Fisher test; c: Non-parametric Mann-Whitney U test; \*: statistically significant; CI: confidence interval; max: maximum; min: minimum; NA: not applied; OR: odds ratio; SD: standard deviation;
<sup>†</sup>: patients with no information were excluded.

The most frequent intravenous prophylactic antibiotic regimen was clindamycin 600mg and gentamicin 240mg (n=28; 62.2%) or clindamycin alone (n=9; 20%) (indicated for less extensive surgeries), with the first dose administered before starting surgery and the last dose up to 48 hours after the start of surgery. Other reported protocols are given in **Suppl. Information**. The mean surgery time was 9 hours, and the mean ICU stay was 3.6 days.

Regarding the main variable of interest in this study, 14/45 patients (31.1%) developed surgical site infection (SSI) (**Tables 1 and 2**). Secretion culture was performed for 3/14 (21.4%) of the patients with SSI, of which one was positive for the alpha-hemolytic *Streptococcus viridans*, while in the cultures of the other two patients no species grew.

**Table 2.**
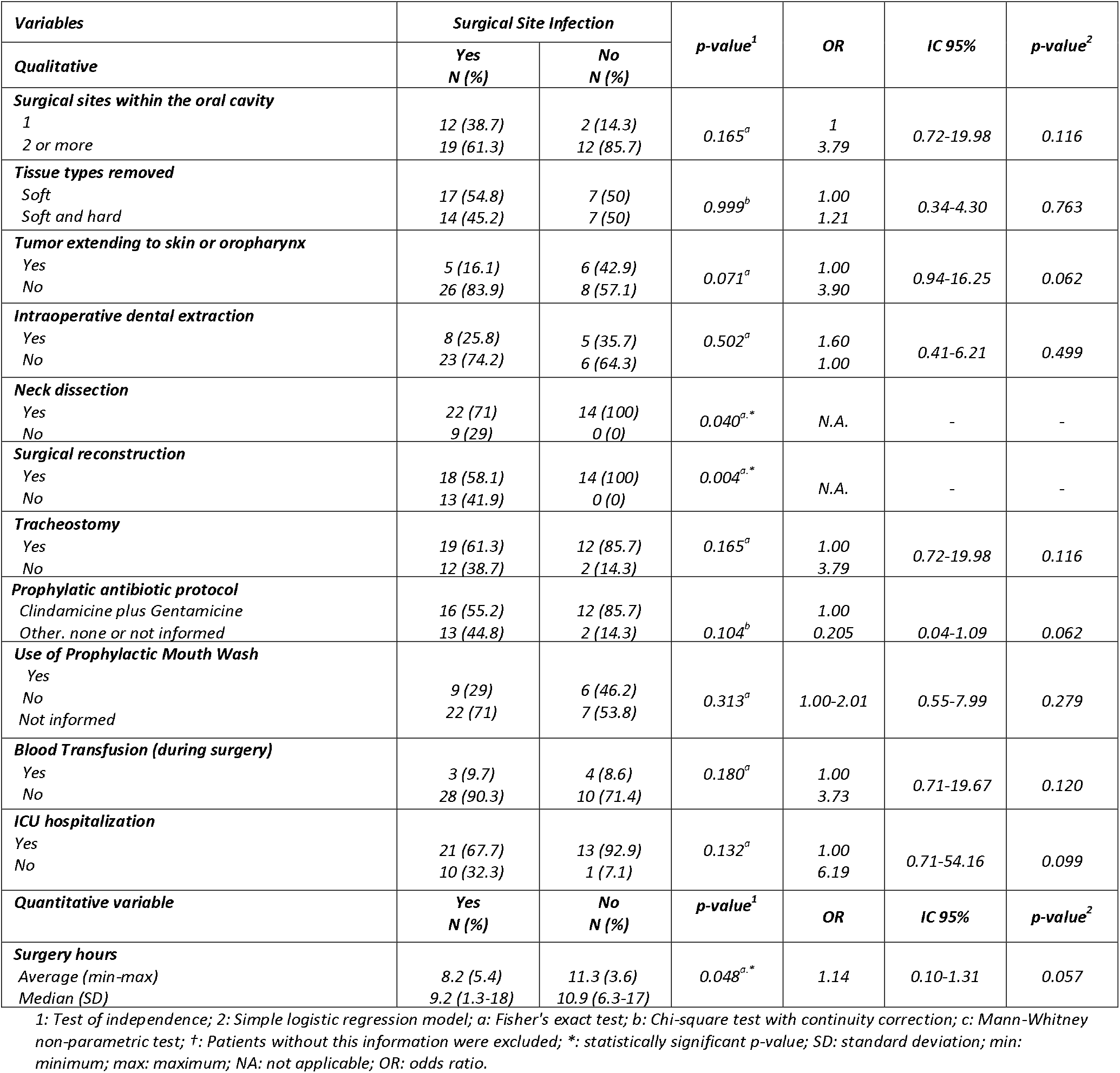
Surgical variables - analysis according to SSI.

| **Variables** | **Surgical Site Infection** |  | **p-value<sup>1</sup>** | **OR** | **IC 95%** | **p-value<sup>2</sup>** |
| --- | --- | --- | --- | --- | --- | --- |
|  | **Yes**<br>**N (%)** | **No**<br>**N (%)** |  |  |  |  |
| **Qualitative** |  |  |  |  |  |  |
| **Surgical sites within the oral cavity** |  |  |  |  |  |  |
| 1 | 12 (38.7) | 2 (14.3) | 0.165<sup>a</sup> | 1<br>3.79 | 0.72-19.98 | 0.116 |
| 2 or more | 19 (61.3) | 12 (85.7) |  |  |  |  |
| **Tissue types removed** |  |  |  |  |  |  |
| Soft | 17 (54.8) | 7 (50) | 0.999<sup>b</sup> | 1.00<br>1.21 | 0.34-4.30 | 0.763 |
| Soft and hard | 14 (45.2) | 7 (50) |  |  |  |  |
| **Tumor extending to skin or oropharynx** |  |  |  |  |  |  |
| Yes | 5 (16.1) | 6 (42.9) | 0.071<sup>a</sup> | 1.00<br>3.90 | 0.94-16.25 | 0.062 |
| No | 26 (83.9) | 8 (57.1) |  |  |  |  |
| **Intraoperative dental extraction** |  |  |  |  |  |  |
| Yes | 8 (25.8) | 5 (35.7) | 0.502<sup>a</sup> | 1.60<br>1.00 | 0.41-6.21 | 0.499 |
| No | 23 (74.2) | 6 (64.3) |  |  |  |  |
| **Neck dissection** |  |  |  |  |  |  |
| Yes | 22 (71) | 14 (100) | 0.040<sup>a,*</sup> | N.A. | - | - |
| No | 9 (29) | 0 (0) |  |  |  |  |
| **Surgical reconstruction** |  |  |  |  |  |  |
| Yes | 18 (58.1) | 14 (100) | 0.004<sup>a,*</sup> | N.A. | - | - |
| No | 13 (41.9) | 0 (0) |  |  |  |  |
| **Tracheostomy** |  |  |  |  |  |  |
| Yes | 19 (61.3) | 12 (85.7) | 0.165<sup>a</sup> | 1.00<br>3.79 | 0.72-19.98 | 0.116 |
| No | 12 (38.7) | 2 (14.3) |  |  |  |  |
| **Prophylactic antibiotic protocol** |  |  |  |  |  |  |
| Clindamicine plus Gentamicine | 16 (55.2) | 12 (85.7) | 0.104<sup>b</sup> | 1.00<br>0.205 | 0.04-1.09 | 0.062 |
| Other, none or not informed | 13 (44.8) | 2 (14.3) |  |  |  |  |
| **Use of Prophylactic Mouth Wash** |  |  |  |  |  |  |
| Yes | 9 (29) | 6 (46.2) | 0.313<sup>a</sup> | 1.00-2.01 | 0.55-7.99 | 0.279 |
| No | 22 (71) | 7 (53.8) |  |  |  |  |
| Not informed |  |  |  |  |  |  |
| **Blood Transfusion (during surgery)** |  |  |  |  |  |  |
| Yes | 3 (9.7) | 4 (8.6) | 0.180<sup>a</sup> | 1.00<br>3.73 | 0.71-19.67 | 0.120 |
| No | 28 (90.3) | 10 (71.4) |  |  |  |  |
| **ICU hospitalization** |  |  |  |  |  |  |
| Yes | 21 (67.7) | 13 (92.9) | 0.132<sup>a</sup> | 1.00<br>6.19 | 0.71-54.16 | 0.099 |
| No | 10 (32.3) | 1 (7.1) |  |  |  |  |
| **Quantitative variable** | **Yes**<br>**N (%)** | **No**<br>**N (%)** | **p-value<sup>1</sup>** | **OR** | **IC 95%** | **p-value<sup>2</sup>** |
| **Surgery hours** |  |  |  |  |  |  |
| Average (min-max) | 8.2 (5.4) | 11.3 (3.6) | 0.048<sup>a,*</sup> | 1.14 | 0.10-1.31 | 0.057 |
| Median (SD) | 9.2 (1.3-18) | 10.9 (6.3-17) |  |  |  |  |

1: Test of independence; 2: Simple logistic regression model; a: Fisher's exact test; b: Chi-square test with continuity correction; c: Mann-Whitney non-parametric test; †: Patients without this information were excluded; \*: statistically significant p-value; SD: standard deviation; min: minimum; max: maximum; NA: not applicable; OR: odds ratio.

We also evaluated the occurrence of SSI according to age. In **Table 3** we observe that younger subjects presented larger and more advanced tumors (T3/T4) as compared to older subjects (median age: 57.2 for T3/T4 x 72.9 years for T1/T2; *p*=0.033).

**Table 3.** Tumor size and clinical stage according to patient age.

| Variables | n | Age |  | p-value<sup>1</sup> |
| --- | --- | --- | --- | --- |
|  |  | Average (SD) | Median (min-max) |  |
| **Tumor size<sup>†</sup>** |  |  |  |  |
| T1/T2 | 18 | 68.19 (14.47) | 72.88 (39.28-88.69) | 0.033* |
| T3/T4 | 20 | 59.42 (10.11) | 57.20 (43.97-81.35) |  |
| **Clinical stage<sup>†</sup>** |  |  |  |  |
| I/II | 14 | 67.25 (15.18) | 69.46 (39.28-88.69) | 0.173 |
| III/IV | 24 | 61.43 (11.29) | 58.80 (43.97-81.35) |  |

<sup>1</sup>: Non-parametric Mann-Whitney U-test.<sup>†</sup>: Patients without this information were excluded from this analysis; \*: indicates statistical significance; SD: Standard deviation.

### Bacterial profiles and SSI

#### Quantitative analysis

Significant variations in the C_T_ values for the 16S-rDNA gene, beta-actin and their ratios **(Suppl. Table 1)** were observed after 170/174 samples (97.7%) were evaluated using average Cycle threshold values (C_T_), and their ratios for 16S-rDNA/human beta-actin according to collection points. In a dependency analysis test, we compared variations across the 4 time points for 148 samples (37 subjects). While the results showed a significant difference across collection points, with reduced bacteria levels after surgery (p<0.01) – no differences were observed when comparing the groups according to SSI occurrence **(Suppl. Figure 1; Suppl. Table 2)**.

#### Qualitative analysis – determining bacterial composition by 16S-rDNA sequencing

Successful amplification of the target gene was achieved for 172/174 samples (98.9%), with an average of 85.352 raw reads/sample (23.013 to 314.457 reads), and an average of 20.205 processed reads/sample, allowing sequencing saturation to be demonstrated **(Suppl. Figure 2)**. Analysis of these 172 samples revealed the presence of 16 phyla, 24 classes, 45 orders, 89 families, 195 genus and 507 bacterial species **(Suppl. Figure 3)**. Taxa originally annotated as *“Ambiguous taxa”, “NA”, “uncultured”, “uncultured bacterium”* and *“unidentified”* were now grouped as unknown genus, totaling 0.9% of all genera in all samples.

Alpha and beta diversities were evaluated after grouping all samples according to collection intervals (I to IV), comparing subjects with or without SSI. At OTU level, for alpha diversity, subjects who developed SSI presented higher bacterial diversity in point IV (12 days after surgery) according to all three indexes used (Observed, Shannon and Simpson; all with *p*<0.05) **(Suppl. Figure 4)**. To evaluate changes in diversity across sampling times, we analyzed the Observed, Shannon, and Simpson indices. Overall, patients with SSI showed a higher bacterial burden and a more even distribution of taxa than those without SSI. In the beta-diversity analysis, the 2D PCoA showed a significant difference at this same time point in the weighted UniFrac analysis (p = 0.043) **(Suppl. Fig. 5)**, suggesting that bacterial composition differs between groups when SSI is present. In summary, at collection 4 there was greater bacterial diversity in patients with established infection. For collection points other than during SSI, diversity profiles were similar and could not predict SSI occurrence. Consistently, the logistic regression model showed no significant association (data not shown), indicating that diversity alone cannot identify patients at higher risk of SSI, since differences were observed only after infection was established.

To better assess changes in diversity across sampling times, we analyzed *Observed*, Shannon, and Simpson indexes. Overall, non-SSI patients showed a greater trend toward a marked post-surgical decrease in bacterial diversity, whereas patients with SSI showed no differences across collections for Shannon and Simpson indexes **(Fig. 2A)**. However, the *Observed* index revealed a significant reduction in OTU richness between the first and third collections **(Fig. 2B)**. This pattern was similar to our previous qPCR analysis of the 16S rDNA V1 region, in which bacterial DNA decreased after surgery. Taken together, both approaches show that oral cancer surgery strongly affects the oral microbiota, reducing both bacterial load and diversity in patients with and without SSI.

**Figure 2.**
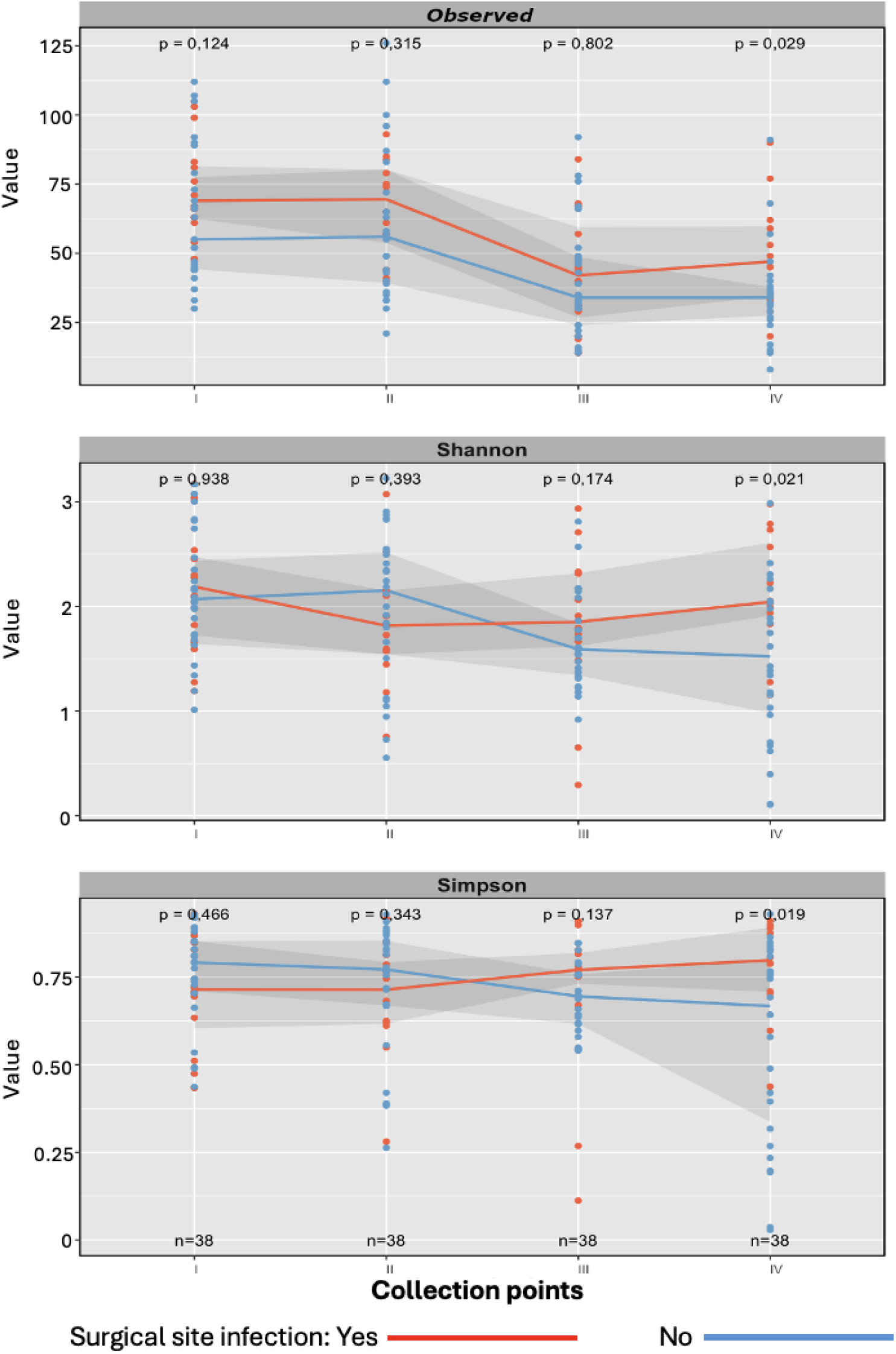
Alpha-diversity values (Observed, Shannon and Simpson) stratified according to the presence (red) or absence (blue) of surgical site infections considering the 4 collection points. Blue and red lines indicate median values; shaded areas indicate 25-75 percentiles; Non-parametric U Mann-Whitney test.

#### Bacterial composition

Of the 194 genera found in the 172 surgical samples, only 12 (6.2%) had an average frequency above 1%, indicating that the oral microbiota is dominated by a few bacterial groups. Adding the frequencies of these 12 genera, they accounted for about 89% of all reads. The remaining 182 identified genera had average frequencies ranging from 0.000058% to 0.98%. **Suppl. Figure 6** shows the inequality among the genera, with a clear dominance of ∼20-25 genera.

A comparison of the top 10 most frequent bacterial genera in patients with and without SSI **(Suppl. Table 3)** shows that, before surgery (collection points 1 and 2), the bacterial profiles were very similar between groups, with only minor differences. However, by time point 3 (post-surgery), a clear change emerged. In patients with no SSI, previously abundant bacteria like *Rothia, Veillonella, Porphyromonas*, and *Fusobacterium* (with a combined sum of 18.9%) dropped sharply in abundance (reaching 2.5% in the combined sum of collection point 3). In the meantime, *Eikenella, Aggregatibacter, Pseudomonas* and *Proteus* increased. By collection point 4 (∼5 days after surgery), the microbiota of non-SSI patients began to return to its pre-surgery status, with *Rothia* and *Veillonella* reappearing as dominant genera. This suggests that patients who do not develop SSI have the capability of an earlier restoring of their pre-operative oral microbiota within about five days after surgery.

A decrease in *Rothia* from collection II to III was observed in both SSI and non-SSI groups, along with a reduction in *Actinobacillus* and *Granulicatella* (which together accounted for 19.58% before surgery but only 0.65% at collection III). Concurrently, *Eikenella, Aggregatibacter*, and *Acinetobacter* increased. However, unlike in patients without SSI, none of the three bacteria that decreased after surgery (*Rothia, Actinobacillus*, and *Granulicatella*) returned to being among the most common genera by the time of SSI (collection IV). Instead, *Staphylococcus*—a known bacterium associated with SSI —appeared on the list along with *Atopium*, suggesting that the development of SSI further alters the bacterial composition. Furthermore, comparing time points 1 and 4 in these patients, only 6 out of the 10 most common genera were the same, and their frequencies differed substantially. For example. among SSI-patients, *Streptococcus* dropped from 39.6% in collection point 1 to just 7.71% in collection point 4. These results indicate that, unlike non-SSI patients, those with infection exhibited a final bacterial profile that was even more divergent from the preoperative baseline, consistent with the notion that infection disrupts the microbiota and prevents restoration of its presurgical profile.

To further assess bacterial fluctuations across collections, the frequency of all 20 genera from **Suppl. Table 3** were analyzed. Bacteria such as *Actinobacillus, Rothia*, and *Veillonella* decreased sharply after surgery, while others like *Eikenella, Pseudomonas* and *Proteus* increased significantly in both groups. *Porphyromonas* increased after surgery in SSI patients while decreasing in non-SSI patients. Finally, *Staphylococcus* grew in the SSI group after surgery, whereas in non-SSI patients it was only detected after surgery (collection point 3), decreasing afterwards. Overall, these findings indicate that major changes in the oral microbiota result from the surgical procedure itself, and these alterations appear to be related to the development of SSI by collection point 4.

In an association analysis, the number of differentially abundant bacteria between patients with and without SSI increased progressively from collections I to IV. Notably, from collection point I to III, the number of genera more abundant in SSI patients increased from 2 to 6, suggesting that surgery may induce oral dysbiosis potentially predisposing to SSI.

Most bacteria significantly abundant in collections I–III were no longer different at collection IV (when infection was established). For example, *Aggregatibacter, Catonella*, and *Solobacterium* stood out only at collection III. This suggests that while these bacteria may contribute to infection onset, their presence triggers dysbiosis that allows other bacteria (e.g. *Prevotella. Porphyromonas. Mogibacterium*) to thrive by the time of SSI. *Morganella* was more frequent in SSI patients across all time points, but its overall abundance was very low (0.02%–15% in only 3 SSI patients) and absent in non-SSI patients, making it unlikely to be necessary for SSI development **(Figure 3)**.

**Figure 3.**
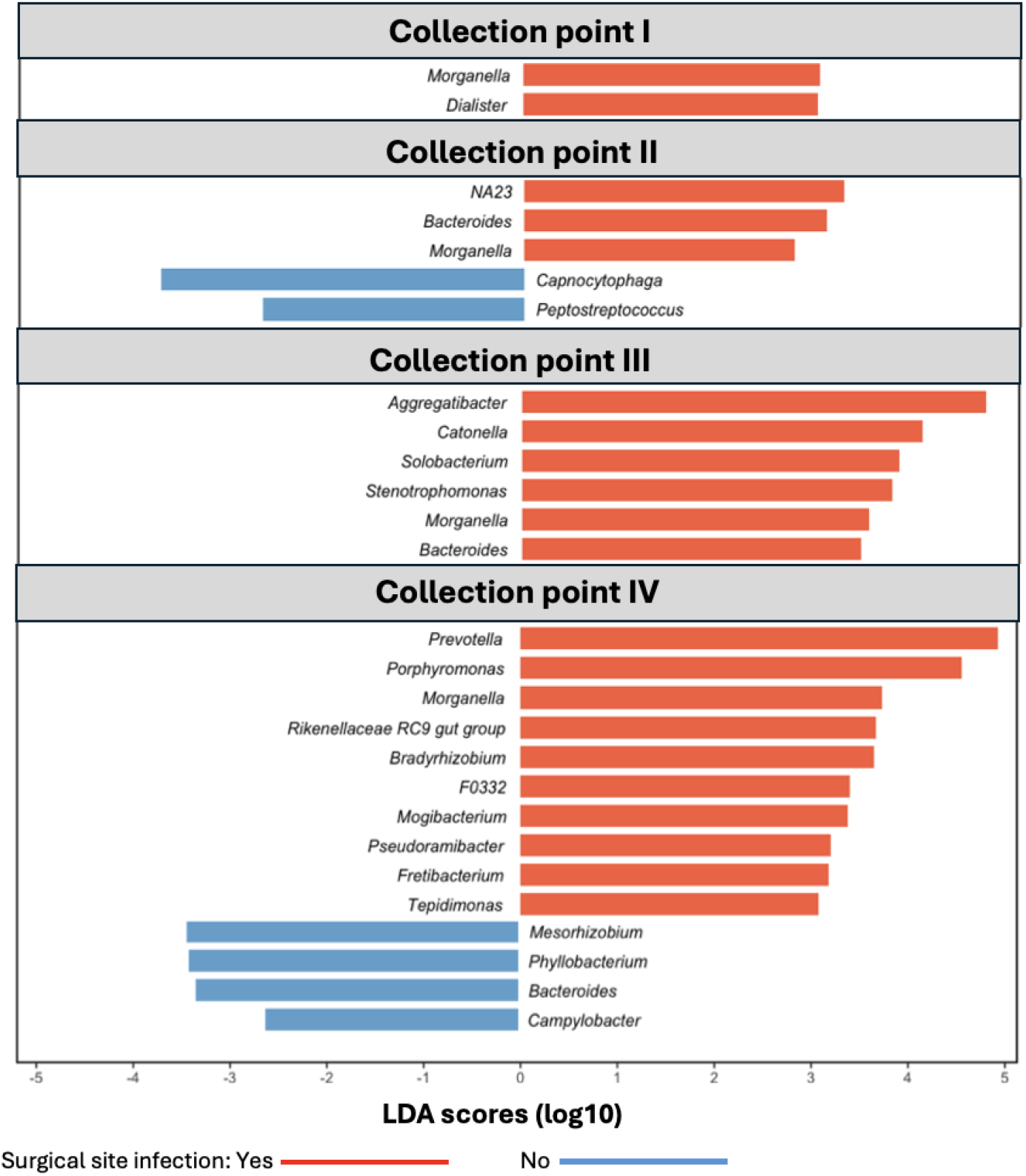
Bacterial genera with differential abundance between patients with and without surgical site infection at each collection time, according to LDA scores (log10) (Mann–Whitney nonparametric test). Obs: bacterium NA23 is an unknown genus within the family Neisseriaceae.

At the time of SSI, 10 genera were more abundant in SSI-positive patients. *Prevotella* and *Porphyromonas* stood out, together accounting for nearly 40% of reads in infected patients vs. 9.45% in non-infected. *Prevotella* was present in all infected cases and *Porphyromonas* in al l but four SSI cases. Both *Prevotella* and *Porphyromonas* possess multiple virulence factors that may exacerbate surgical site infections and impair healing.

The other 8 genera represented only 0.89% of sequences, making their actual role unclear. In non-infected patients, the more abundant bacteria (e.g.. *Bacteroides*) represented only 0.62%, suggesting no protective role. Thus, *Prevotella* and *Porphyromonas* consistently prevailed at SSI, though they likely are not the primary cause but rather secondary consequences of earlier dysbiosis (e.g. by an increment of *Aggregatibacter* at collection point III). Common culture-based SSI pathogens like *Pseudomonas* and *Staphylococcus* were not significantly associated with infection in this study, possibly due to genus-level analysis or sporadic occurrence in this population.

In logistic regression analysis, *Aggregatibacter* at collection III (post-surgery. up to 5 days) was a significant predictor. Each 1% increase raised SSI odds by 10% (p=0.012) (Table 4). Frequencies above 0.044% were associated with a 5.67-fold higher chance of SSI (p=0.024), suggesting *Aggregatibacter* as a potential risk indicator.

**Table 4.**
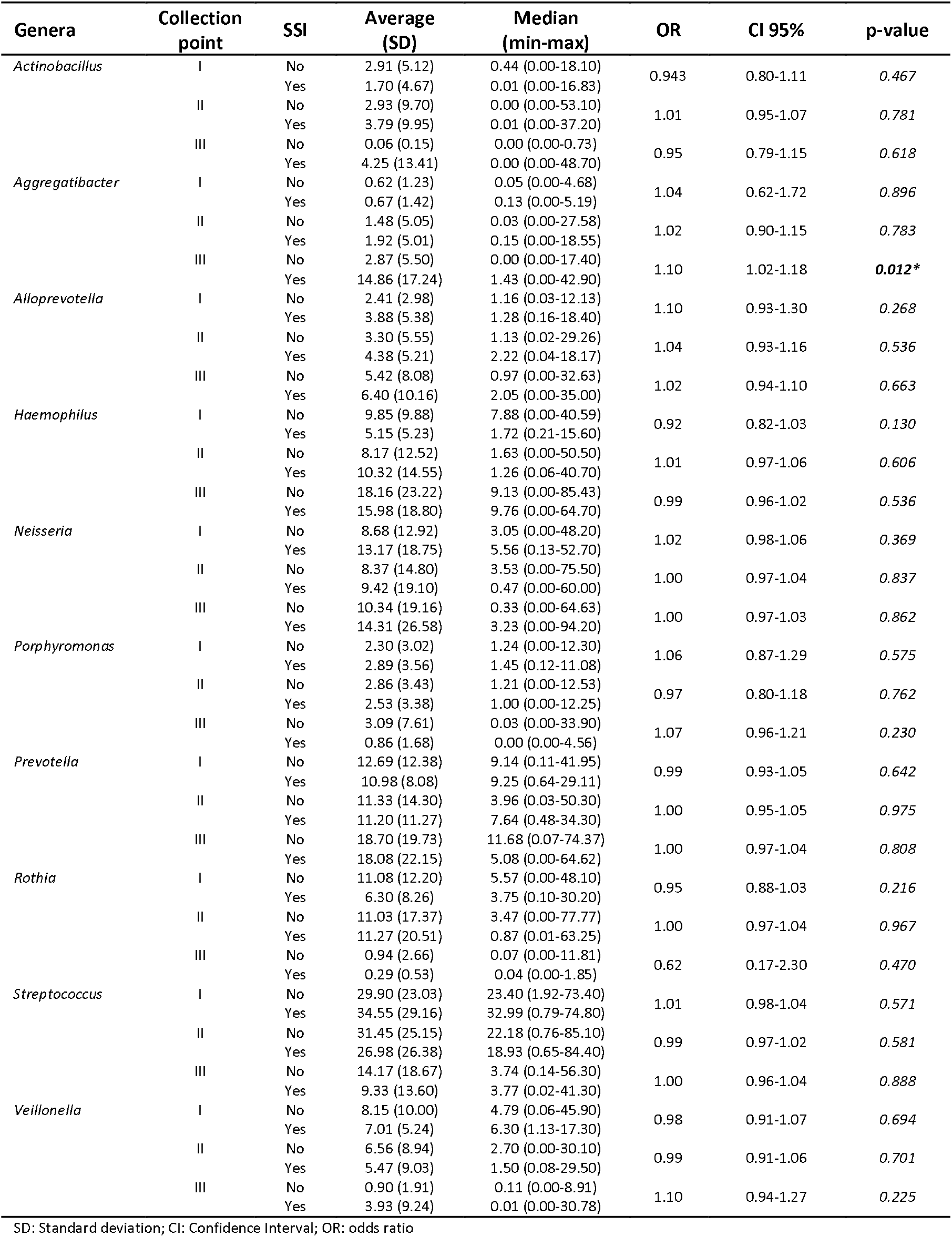
Logistic regression model of the 10 most frequent genera in relation to collection time and presence of surgical site infection.

## Discussion

The quantitative variations in bacteria, observed as crude median Ct values for 16S gene, as well as beta-actin/16S ratios are likely due to asepsis procedures performed at the peri-surgical interval and antibiotic use impacting the microbiota (Gallant, 2024) as well as the removal of human tissues and debris, the surgical procedure itself, and the following wound healing that attract cells relevant to this process and increase human DNA amounts after surgery (Dohmen et al., 2007). Peri-operative topical antiseptic protocols, including povidone-iodine and chlorhexidine gluconate, have been shown by Zenga et al. (2022) to significantly decrease oral bacterial bioburden from pre-operative levels. Additionally, prophylactic intravenous antibiotics administered for 24 hours post-operatively—such as ampicillin/sulbactam or clindamycin with gentamicin—further modulate the oral microbial community, as demonstrated by Yang et al. (2014). Oral cancer surgery itself also involves violation of the oral mucosa with spillage of the oral microbiome into normally sterile neck tissues, contributing to post-operative dysbiosis (Gallant, 2024). The combined effect of these interventions and surgical trauma results in measurable shifts human:bacteria ratios, bacterial load and community composition during the peri-operative period.

Advanced clinical stage, cervical dissection, surgical reconstruction, and longer operative time are established risk factors for SSI in oral cancer patients. Karakida et al. (2010) identified advanced T-stage and longer surgery duration as significant predictors. Chen et al. (2025) confirmed that flap reconstruction (OR 4.5) and mandibulectomy (OR 4.4) strongly increase SSI risk. Qin et al. (2026) demonstrated that neck dissection and prolonged operative time are key associated factors, while Lee et al. (2011) identified oral cavity tumor location as an additional independent risk factor.

Regarding age, studies report that SSI is more frequent in older patients, as they generally have poorer immune status, making them more susceptible to infections (Esposito, 2001). However, our study revealed the opposite, with younger patients presenting higher incidence of SSI. Nevertheless, through association analysis we observed that, in general, the younger patients had more advanced and larger tumors (T3/T4) as compared to elderly patients (p=0.033) (Table 3), as shown by others (Musella et al., 2026), leading to longer and more aggressive surgical treatment that would facilitate the occurrence of SSI.

Similar to our findings, a study conducted by Sun et al. 2017, found that out of 163 genera in tongue samples collected with swabs, the 15 most abundant (9.2%) represented between 70.2% and 82% of all reads in each sample. This shows that despite the large number of bacterial genera in the human oral cavity, very few predominate and represent most of the bacterial mass in the oral cavity; in our study, only 12 genera above 1% were found out of a total of 194 **(Suppl. Figure 3)**. Due to their higher frequency, we believe these are the bacteria that have the most impactful effect on the oral microbiota and thus on the oral cavity. Bacteria in large quantities likely occupy ecological niches, controlling the establishment and growth of other microorganisms, especially those present in low frequency. However, in dysbiosis events caused by interventions, medications or other factors, this dynamic can be altered.

Considering only the bacterial present before surgery, few differences were seen between SSI and non-SSI groups and the microbiota was stable for both patients groups between collection points I and II **(Suppl Table 3; Suppl. Figure 2)**. This reinforces the stability of the oral microbiota in this average 29 days of interval, in the absence of interventions, as demonstrated by others (Yang et al.. 2018; Belstrøm et al.. 2016; Costello et al.. 2009; Lundtorp-Olsen et al., 2021). Whereas the microbiota pattern was stable before surgery, a clear altered pattern was seen afterwards. In patients with no SSI, previously abundant bacteria like *Rothia, Veillonella, Porphyromonas*, and *Fusobacterium* (with a combined sum of 18.9%) dropped sharply in abundance (reaching 2.5% in the combined sum of collection point 3). In the meantime, *Eikenella, Aggregatibacter, Pseudomonas* and *Proteus* increased. By collection point 4 (∼5 days after surgery), the microbiota of non-SSI patients began to return to its pre-surgery status, with *Rothia* and *Veillonella* reappearing as dominant genera. This suggests that patients who do not develop SSI have the capability of an earlier restoring of their pre-operative oral microbiota within about five days after surgery.

Specific bacterial species with increased frequency in the SSI group, including *Porphyromonas* and *Prevotella*, have been implicated in modulating host cell signaling pathways and creating a microenvironment conducive to infection (Santacroce et al., 2025; Qin et al., 2024). Furthermore, polymicrobial infections—predominantly involving *Staphylococcus aureus* and *Pseudomonas aeruginosa*—have been documented as the leading microbiological trend in oral cavity carcinoma SSI (Cirignaco et al., 2025).

*Prevotella* and *Porphyromonas* possess multiple virulence factors that may exacerbate surgical site infections and impair healing. Botta et al. (1994) and Marre et al. (2026) demonstrated that these organisms express adhesins and capsules involved in adhesion and abscess formation, being capable of degrading matrix proteins and to activate plasminogen, indicating tissue-destroying capabilities. *Porphyromonas gingivalis* secretes gingipains that degrade key cell-cell junction proteins. Katz et al. (2002) demonstrated that lysine-specific gingipain hydrolyzes E-cadherin in epithelial monolayers. Sheets et al. (2005) showed that gingipains cleave N-cadherin and VE-cadherin in endothelial cells, leading to loss of adhesion. Takeuchi et al. (2019) found that gingipains degrade JAM-1, increasing epithelial permeability. Golubkov et al. (2020) provided proteomic evidence that most gingipain substrates are adhesion molecules, causing cell detachment. Farrugia et al. (2020) demonstrated gingipain-dependent degradation of PECAM-1 and VE-cadherin in vivo. Hajishengallis et al. (2012) detailed anti-phagocytic mechanisms including complement degradation and intracellular survival pathways. Sharma et al. (2022) documented additional virulence factors in *Prevotella* spp., including proteases and secretion systems. Accordingly, Brook (2002) identified pigmented *Prevotella* and *Porphyromonas* as the most frequently recovered organisms from post-surgical wounds after head and neck cancer surgery.

*Aggregatibacter*, a genus that increased in the SSI group immediately after surgery, is an opportunistic pathogen linked to various oral and systemic infections. In periodontal disease, it is an early colonizer that tolerates oxygen and hydrogen peroxide, but after establishing itself and releasing pathogenic factors, it is often replaced by more strict anaerobes (Moustafa et al., 2021). Under anaerobic, short-chain fatty acid-rich conditions similar to those found in deep periodontal pockets, *Aggregatibacter* strains upregulate genes involved in virulence, colonization, and immune evasion, forming robust biofilms that alter the local microenvironment (Moustafa et al., 2021). This ecological behavior is consistent with the **“keystone pathogen”** hypothesis, whereby a low-abundance pioneer species can reshape the microbial community and drive disease by manipulating host defenses (Hajishengallis et al., 2012). Thus, *Aggregatibacter*’s virulence mechanisms promote a dysbiotic environment conducive to infection, potentially benefiting the obligate anaerobic bacteria—*Prevotella and Porphyromonas -* that later predominate at the time of SSI.

In our study, an *Aggregatibacter* frequency exceeding 0.044% was associated with 5.67-fold increased odds of developing SSI, suggesting that this bacterium plays a catalytic role in infection during the immediate postoperative period. However, its relative abundance diminished by the time of established infection (collection IV). This temporal pattern is consistent with ecological principles of microbial succession observed in other polymicrobial infections, where early colonizers termed “founder species” can biochemically remodel the local microenvironment, effectively “preparing the soil” for secondary opportunistic pathogens (Srinivasan et al., 2023). We hypothesize that *Aggregatibacter* acts as such a keystone pioneer, altering the host environment (e.g., oxygen tension, nutrient availability) to favor the outgrowth of obligate anaerobes, specifically *Prevotella* and *Porphyromonas*, which dominated at the clinical onset of SSI. This successional pattern aligns with observations in chronic wounds, where an increase in anaerobic populations occurs as infections progress and mature (Srinivasan et al., 2023; Coluccio et al., 2024). While the ecological principle of microbial succession has been described in other chronic infections (Srinivasan et al., 2023; Farrell et al., 2025), this study is among the first to empirically demonstrate a specific “founder-effect” cascade—from *Aggregatibacter* to obligate anaerobes—predicting SSI in oral cancer patients. This mechanistic insight suggests that early postoperative interventions targeting at-risk patients—such as extending or adjusting prophylactic antibiotic regimens—could disrupt this successional cascade, potentially reducing hospital stay, healthcare costs, and long-term functional loss.

To our knowledge, only one previous study attempted to identify head and neck cancer surgery patients at higher SSI risk via microbiota, but it was limited to 13 bacterial species and only counted growth. Our sequencing approach identified specific bacterial fluctuations before and after surgery and their association with SSI. Limitations include a small patient sample (precluding multiple logistic regression), but the large set of 172 samples evaluated here, collected longitudinally before and after surgery, enabled extensive metagenomic analysis of the microbiota in multiple time points allowing the tracing microbial profiles and identifying a potential bacterial indicator for SSI risk.

## Data Availability

All data produced in the present study are available upon reasonable request to the authors

## Acknowledgements

The authors gratefully acknowledge the financial support of the Science and Technology Department of the Ministry of Health, Brazil (DECIT, PRONON, SIPAR 2500.035-167/2015-23; PI: ED-N). This study was financed in part by the Coordenação de Aperfeiçoamento de Pessoal de Nível Superior – Brasil (CAPES) – Finance Code 001, which provided a doctoral scholarship to MSS.

