## Supplemental information, tables and figures for "Dynamic Shifts in the Oral Microbiota Following Cancer Surgery: A 172-Sample Longitudinal Study of Surgical Site Infection Risk"

**Suppl. Info SSI_Marianna**

Intravenous prophylactic antibiotic regimens - Besides the treatment protocols indicated in the main text, some patients received other regimens such as clindamycin and ceftriaxone (n=2; 4.4%), clindamycin and garamicin (n=1; 2.2%), clindamycin and ciprofloxacin (n=1; 2.2%), clindamycin and rocefin (n=1; 2.2% – as indicated for a patient diagnosed with tuberculosis who was already using rocefin), as well as tazocin (n=1; 2.2% – indicated for an extensive surgery performed in two operative stages).

**Suppl. Table 1** – Average $C_T$ values for the target genes according to collection points.

| Target genes & collection points | n | Average $C_T$ value (SD) | Median $C_T$ value (min-max) | p-value[1] |
|---|---|---|---|---|
| **16S-V1** | | | | |
| I | 37 | 18.07 (2.08) | 18.41 (13.80-21.70) | *<0.001** |
| II | 37 | 19.10 (3.14) | 19.02 (14.46-27.02) | |
| III | 37 | 21.42 (3.40) | 21.63 (16.25-30.96) | |
| IV | 37 | 22.78 (3.73) | 22.53 (16.76-32.25) | |
| **Human beta-actin** | | | | |
| I | 37 | 28.65 (1.95) | 27.94 (25.80-33.17) | *<0.001** |
| II | 37 | 28.57 (1.59) | 28.51 (25.96-33.31) | |
| III | 37 | 28.07 (2.34) | 27.83 (25.41-38.49) | |
| IV | 37 | 27.62 (1.31) | 27.45 (25.31-30.41) | |
| **$C_T$-ratio (Human beta-actin/16S-V1)** | | | | |
| I | 37 | 1.60 (0.18) | 1.60 (1.29-2.10) | *<0.001** |
| II | 37 | 1.53 (0.21) | 1.51 (1.04-2.01) | |
| III | 37 | 1.33 (0.17) | 1.30 (0.99-1.63) | |
| IV | 37 | 1.24 (0.19) | 1.26 (0.84-1.68) | |

[1]: Non-parametric Friedman test; p-value was calculated across collection points for each gene; *:indicates statistically significant p-values; SD: Standard deviation.

**Suppl. Table 2** – Average Cт values for the target genes according to collection time point and presence/absence of SSI.

| Time point | SSI | n | Average (SD) | Median (min-max) | p-value[1] | OR | CI 95% | p-value[2] |
|---|---|---|---|---|---|---|---|---|
| **Cт for 16S-V1** | | | | | | | | |
| I | No | 29 | 18.42 (2.29) | 18.50 (13.80-24.27) | | | | |
| | Yes | 13 | 18.18 (2.20) | 18.65 (14.72-21.70) | 0.881 | 0.95 | 0.71-1.28 | 0.746 |
| II | No | 30 | 19.21 (2.99) | 19.26 (14.46-25.95) | | | | |
| | Yes | 14 | 19.54 (3.42) | 18.36 (15.55-27.02) | 0.940 | 1.04 | 0.84-1.27 | 0.732 |
| III | No | 27 | 22.15 (3.77) | 22.01 (16.25-30.96) | | | | |
| | Yes | 13 | 20.51 (2.19) | 20.74 (16.65-22.69) | 0.266 | 0.85 | 0.68-1.06 | 0.157 |
| IV | No | 30 | 22.28 (4.11) | 21.24 (16.76-32.48) | | | | |
| | Yes | 14 | 24.36 (3.56) | 23.40 (17.74-30.49) | 0.062 | | | |
| **Cт for human beta-actin** | | | | | | | | |
| I | No | 29 | 28.33 (1.61) | 27.94 (25.80-33.17) | | | | |
| | Yes | 13 | 29.25 (2.39) | 28.70 (25.94-33.05) | 0.348 | 1.29 | 0.91-1.83 | 0.154 |
| II | No | 30 | 28.37 (1.24) | 28.29 (25.96-31.13) | | | | |
| | Yes | 14 | 28.94 (1.94) | 28.89 (26.43-33.31) | 0.420 | 1.29 | 0.84-1.99 | 0.250 |
| III | No | 27 | 28.12 (2.71) | 27.39 (25.41-38.49) | | | | |
| | Yes | 13 | 27.90 (1.06) | 28.04 (25.93-29.45) | 0.554 | 0.96 | 0.70-1.30 | 0.773 |
| IV | No | 30 | 27.49 (1.34) | 27.29 (25.31-30.41) | | | | |
| | Yes | 14 | 27.99 (1.03) | 27.76 (26.11-29.89) | 0.158 | | | |
| **Cт ratio (human beta-actin/16S-V1)** | | | | | | | | |
| I | No | 29 | 1.56 (0.16) | 1.50 (1.26-1.93) | | | | |
| | Yes | 13 | 1.63 (0.23) | 1.62 (1.29-2.10) | 0.348 | 1.02 | 0.99-1.06 | 0.221 |
| II | No | 30 | 1.51 (0.22) | 1.46 (1.04-2.01) | | | | |
| | Yes | 14 | 1.51 (0.21) | 1.52 (1.10-1.82) | 0.753 | 1.00 | 0.97-1.03 | 0.971 |
| III | No | 27 | 1.29 (0.18) | 1.30 (0.99-1.59) | | | | |
| | Yes | 13 | 1.37 (0.15) | 1.34 (1.17-1.63) | 0.199 | 1.03 | 0.99-1.07 | 0.170 |
| IV | No | 30 | 1.27 (0.21) | 1.29 (0.81-1.68) | | | | |
| | Yes | 14 | 1.17 (0.18) | 1.20 (0.91-1.59) | 0.082 | | | |

[1]: Non-parametric Mann-Whitney U test. [2]: Simple logistic regression model; SD: Standard deviation; SSI: Surgical Site Infection; OR: odds ratio.

**Suppl. Table 3** – The top 10 most abundant genera according to collection point, for subjects with and without SSI.

| Top 10 | No SSI | | SSI | |
|---|---|---|---|---|
| | Genera | Frequency (%) | Genera | Frequency (%) |
| **Ranking collection point I** | | | | |
| 1 | *Streptococcus* | 31.93 | *Streptococcus* | 39.6 |
| 2 | *Prevotella* | 12.23 | *Neisseria* | 13.84 |
| 3 | *Rothia* | 11.56 | *Prevotella* | 9.03 |
| 4 | *Neisseria* | 9.62 | *Rothia* | 7.34 |
| 5 | *Haemophilus* | 8.94 | *Veillonella* | 6.02 |
| 6 | *Veillonella* | 7.84 | *Haemophilus* | 5.62 |
| 7 | *Actinobacillus* | 3.31 | *Porphyromonas* | 2.85 |
| 8 | *Porphyromonas* | 2.19 | *Alloprevotella* | 2.64 |
| 9 | *Alloprevotella* | 1.95 | *Fusobacterium* | 1.74 |
| 10 | *Granulicatella* | 1.31 | *Actinobacillus* | 1.44 |
| **Ranking collection point II** | | | | |
| 1 | *Streptococcus* | 38.58 | *Streptococcus* | 29.33 |
| 2 | *Prevotella* | 9.78 | *Rothia* | 12.05 |
| 3 | *Rothia* | 9.04 | *Prevotella* | 11.04 |
| 4 | *Neisseria* | 8.55 | *Haemophilus* | 10.83 |
| 5 | *Haemophilus* | 7.36 | *Neisseria* | 8.55 |
| 6 | *Veillonella* | 5.83 | *Actinobacillus* | 5.39 |
| 7 | *Actinobacillus* | 4.15 | *Veillonella* | 4.91 |
| 8 | *Alloprevotella* | 2.52 | *Alloprevotella* | 3.95 |
| 9 | *Porphyromonas* | 2.15 | *Granulicatella* | 2.14 |
| 10 | *Fusobacterium* | 1.92 | *Porphyromonas* | 1.94 |
| **Ranking collection point III** | | | | |
| 1 | *Haemophilus* | 18.61 | *Neisseria* | 26.8 |
| 2 | *Streptococcus* | 16.97 | *Prevotella* | 18.84 |
| 3 | *Prevotella* | 15.40 | *Aggregatibacter* | 14.26 |
| 4 | *Neisseria* | 14.78 | *Streptococcus* | 11.33 |
| 5 | *Alloprevotella* | 5.46 | *Haemophilus* | 10.74 |
| 6 | *Eikenella* | 3.62 | *Alloprevotella* | 6.00 |
| 7 | *Aggregatibacter* | 3.07 | *Porphyromonas* | 3.47 |
| 8 | *Pseudomonas* | 2.57 | *Veillonella* | 2.18 |
| 9 | *Proteus* | 2.47 | *Acinetobacter* | 1.02 |
| 10 | *Actinobacillus* | 2.43 | *Eikenella* | 0.72 |
| **Ranking collection point IV** | | | | |
| 1 | *Neisseria* | 26.87 | *Prevotella* | 31.69 |
| 2 | *Streptococcus* | 20.10 | *Streptococcus* | 7.71 |
| 3 | *Rothia* | 11.77 | *Aggregatibacter* | 7.48 |
| 4 | *Prevotella* | 8.91 | *Porphyromonas* | 7.48 |
| 5 | *Haemophilus* | 7.28 | *Neisseria* | 6.26 |
| 6 | *Aggregatibacter* | 3.80 | *Veillonella* | 4.69 |
| 7 | *Veillonella* | 3.49 | *Atopobium* | 3.23 |
| 8 | *Alloprevotella* | 3.40 | *Staphylococcus* | 2.82 |
| 9 | *Corynebacterium* | 1.54 | *Alloprevotella* | 2.64 |
| 10 | NA7 | 1.20 | *Haemophilus* | 2.64 |

SSI: Surgical Site Infection

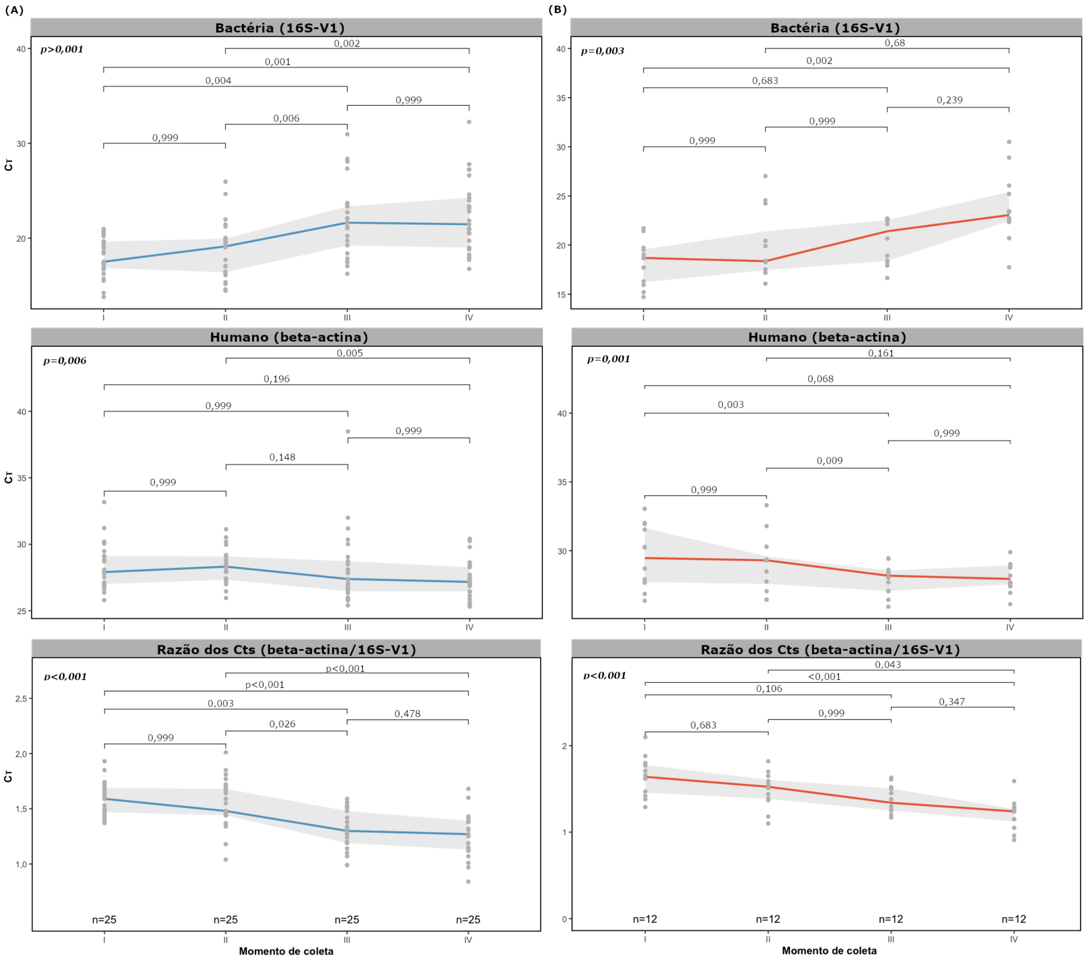


**Suppl. Figure 1** – Representation of CT values (16S for bacteria. beta-actin for human and their ratios) according to collection points (I to IV. **Figure 1**). Panel A (blue lines) indicates CT values for subjects with no SSI and Panel B (red lines) indicates CT values for subjects with SSI. Colored continuous lines represent median values and shadowed areas indicate 25-75 percentiles; Non-parametric Friedman test).

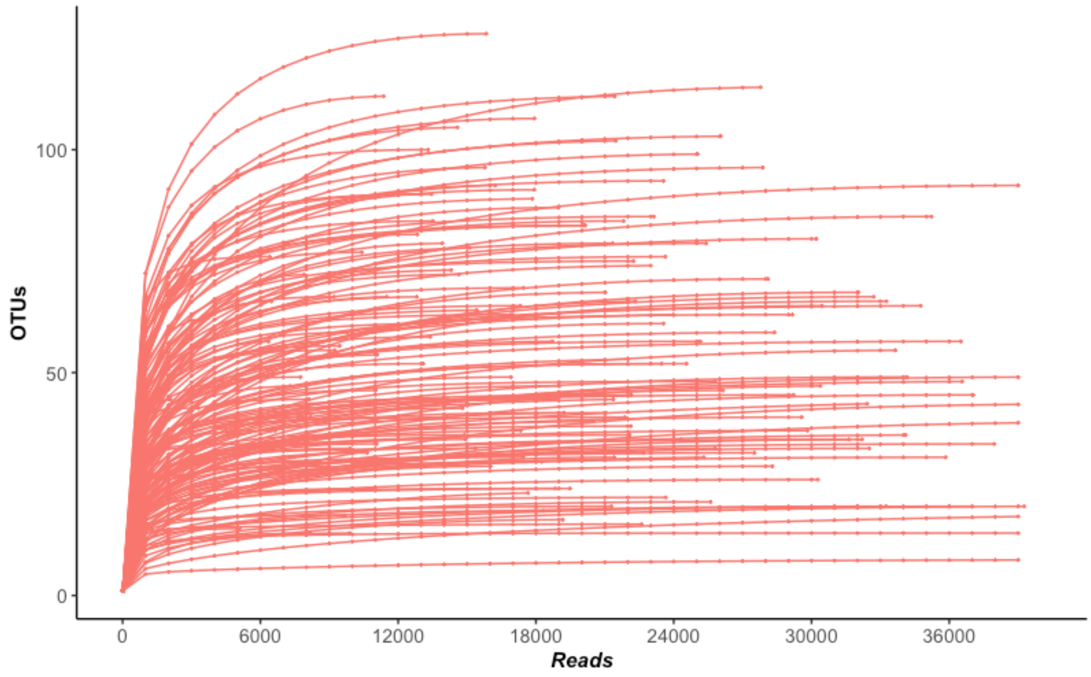


**Suppl. Figure 2** – Saturation curve OTUs after the sequencing of 172 samples.

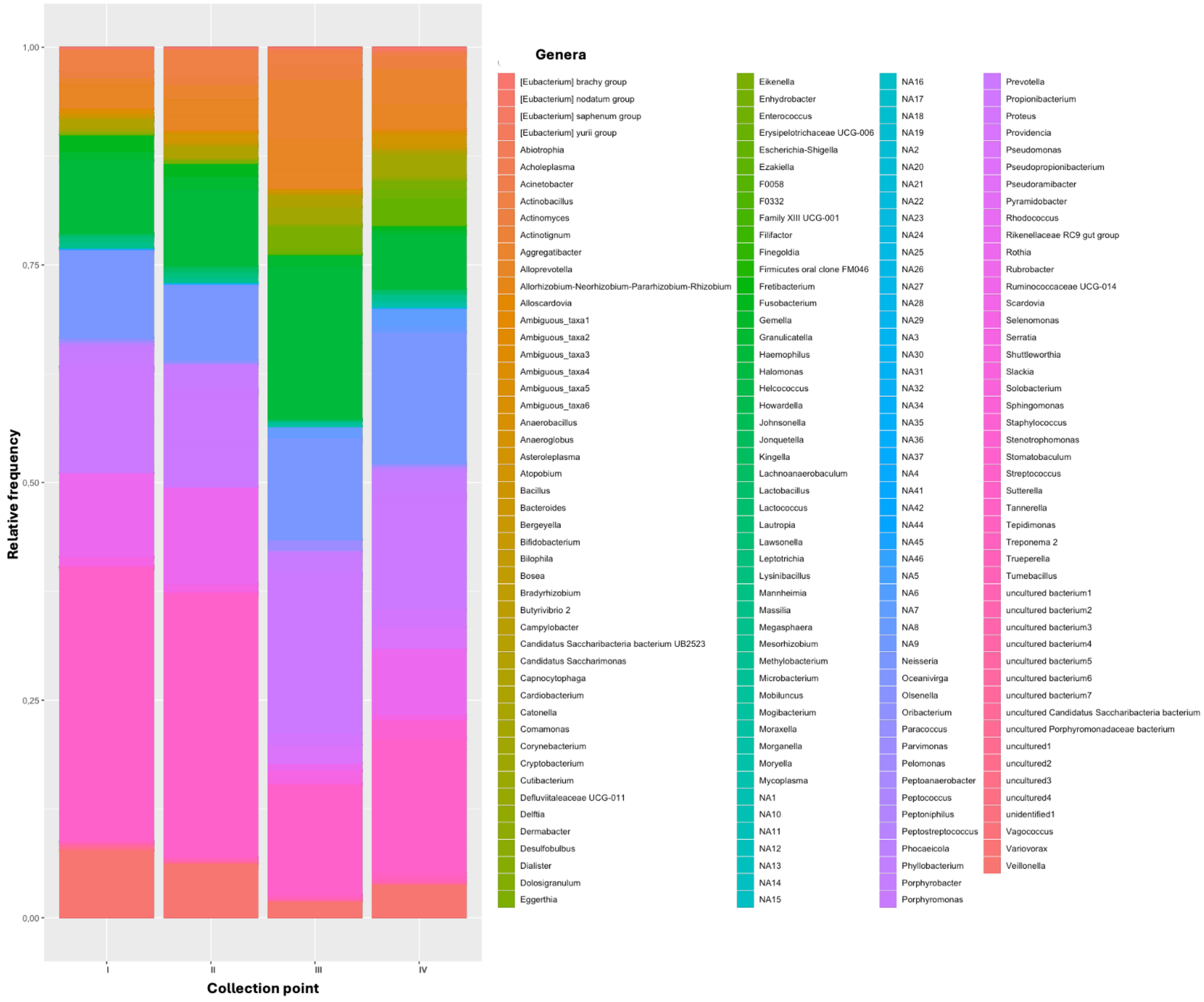


**Suppl. Figure 3** – Relative Frequency of bacteria (according to genera), from collection points I to IV.

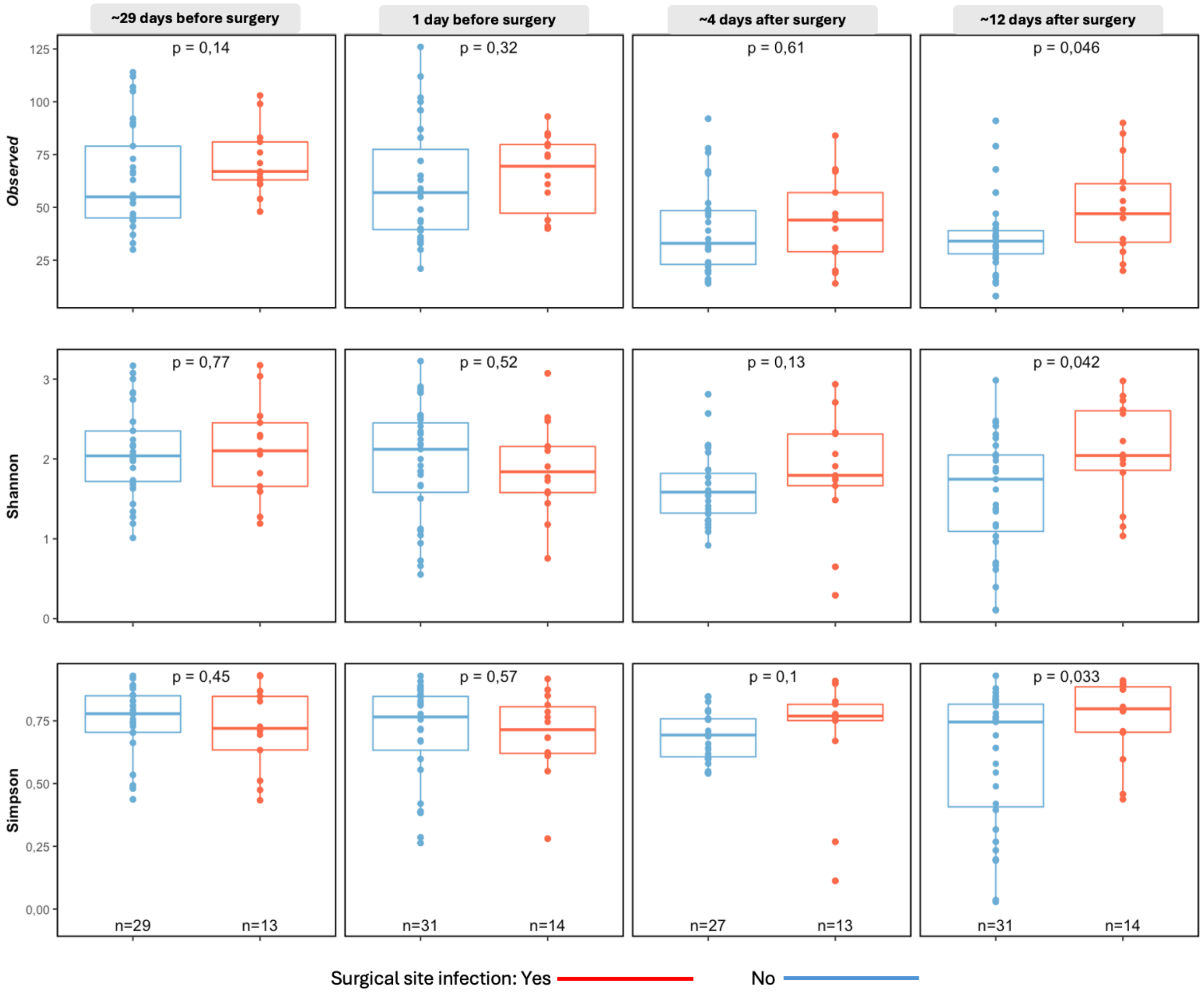


**Suppl. Figure 4** – Alpha diversity according to presence or absence of Surgical Site Infections (SSI) in the 4 collection points (non-parametric Mann-Whitney U-test).

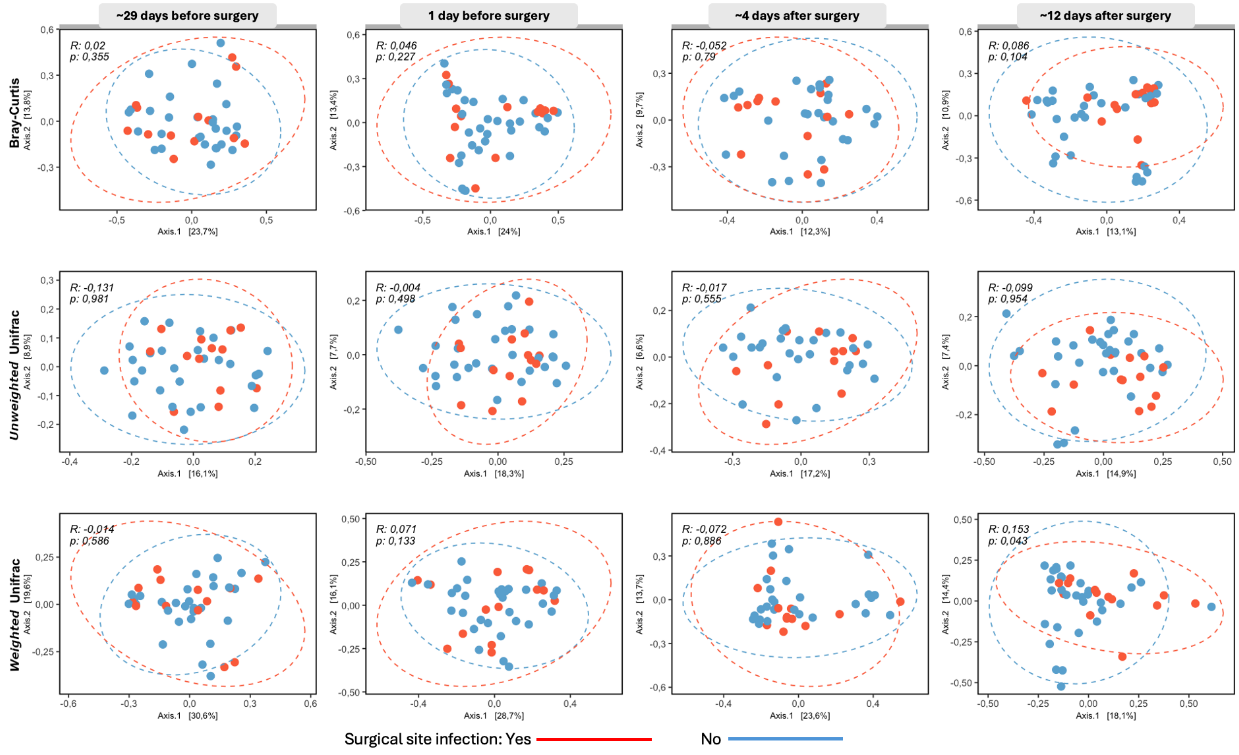


**Suppl. Figure 5** – Beta diversity according to presence or absence of Surgical Site Infections (SSI) in the 4 collection points (*R*: ANOSIM similarity coefficient).

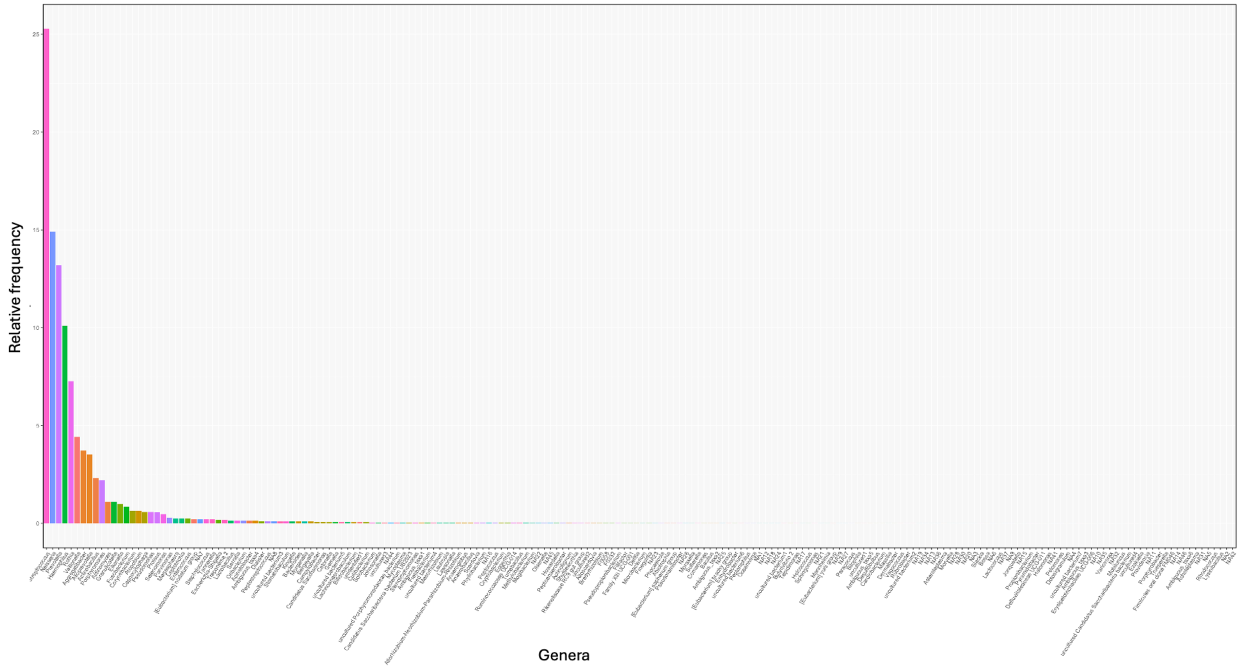


**Suppl. Figure 6** – Relative frequency of the 194 bacteria genera found in the 172 samples sequenced.
